# Use of the Ages and stages questionnaire in Bradford as a measure of Child Development (ABCD): a mixed methods study protocol

**DOI:** 10.1101/2025.08.14.25333745

**Authors:** Hollie C. Henderson, Kate E. Mooney, Kate Morton, Lois Armour, Dawn Lee, Sara Ahern, Sarah L. Blower, Josie Dickerson

**Author notes:** **Corresponding author:** Hollie C Henderson.

## Abstract

The Ages and Stages Questionnaire® is the earliest routinely recorded measure of children’s development at age two, administered in England. However, there have been few studies on the acceptability of this measure within a UK setting, especially for families at risk of inequity. To ensure all children and families receive the necessary support, a reliable measure of child development is essential. This mixed-methods study aims to address the current evidence gaps by investigating if the Ages and Stages Questionnaire® is acceptable, implementable, and equitable for use by health professionals and families as an overall measure of child development in a UK setting.

This study will use data from Born in Bradford’s Better Start birth cohort, which recruits from three ethnically diverse, socioeconomically deprived inner-city wards of the Bradford District. Rich baseline sociodemographic information will be linked to individual-level local health visiting data to describe how many children have records of an Ages and Stages Questionnaire®, investigate the factors associated with children having a record and describe differences in questionnaire scores by sociodemographic characteristics. To support interpretation of the quantitative analyses within the local context, qualitative semi-structured interviews will be conducted to explore health professionals’ perspectives and experiences of implementing the Ages and Stages Questionnaire® in Bradford. We will analyse the interview data using descriptive thematic analysis and integrate the data using the process of triangulation. We will use the Health Equity Implementation Framework to underpin our approach.

This study has been designed with the Bradford Health Visiting Service to support development of locally relevant, evidence-based recommendations for improving the implementation of the Ages and Stages Questionnaire®. The findings from this research will also provide crucial insights into the use of the Ages and Stages Questionnaire® as a tool for measuring early child development, particularly in ethnically diverse, urban populations.

## Introduction

The early years of life, from when a child is conceived up to the age of four, is a critical period for child development [1]. Early identification of developmental concerns, followed by intervention and additional support to parents, are most effective at preventing problems later in life [2]. In England, the 0-to-5 Healthy Child Programme (HCP), delivered universally by health visiting teams via a ‘proportionate universal’ approach, provides support for all families, with additional support for those who need it the most [3]. The HCP requires local authorities to commission the delivery of a minimum of five health visiting contacts for every child and family in England before the child is five years old. The five statutory contacts are: an antenatal visit after 28 weeks of pregnancy; a new birth visit within 14 days; at 6-8 weeks; at 9-12 months; and at 2-2.5 years.

As part of the 2-2.5 year review, the Ages and Stages Questionnaire® Third Edition (ASQ-3™; Hereon, ASQ-3) is used, which assesses child development across five domains: communication, gross motor skills, fine motor skills, problem solving, and personal-social development [4]. NHS England stipulates that parents are sent the ASQ-3 two weeks before their 2-2.5 year review and asked to complete the questionnaire with their child prior to their appointment. This allows parents to try out some of the activities covered by the questionnaire with their child in an environment that is comfortable and familiar to the child. During the 2-2.5 year review, a health professional will review the ASQ-3 with the parents [5]. The Ages and Stages Questionnaires®: Social-Emotional health-2 (ASQ®: SE-2, Hereon, ASQ: SE-2), is an additional set of questions about behavioural and social-emotional development in young children, and is optional as part of 2-2.5 year review [6]. However, in some areas of England, local guidance advises that the ASQ: SE-2 should always be used as part of this review.

The ASQ-3 was originally developed in the United States by Squires and colleagues [4]. Several studies in the United States and other parts of the world have shown this to be a validated screening tool to identify developmental concerns in an individual child [7]. The Department for Health and Social Care implemented the ASQ-3 as a population health measure to help monitor child development across England from year to year, not as an individual assessment or screening tool. It was identified as suitable for national use as part of the HCP 2-2.5 year reviews following a review of child development measures, though it was noted at the time that the ASQ-3 had not been validated or standardised for use in England [8] [9]. The national license for ASQ-3 also does not cover digital use of the tool or languages other than British English [10].

In practice, however, the guidance for use of the ASQ-3 in England is unclear. For example, Public Health England (the commissioners of the HCP services) state that the universal 2-2.5 year review provides an opportunity to identify children who are not developing as expected and require additional early support [11]. However, researchers have suggested that due to lack of validation of the ASQ-3 in England, the ASQ-3 should only be used as a population measure of child development for monitoring and to inform interventions, not as an individual assessment or screening tool [12, 13].

Despite the ASQ-3 being routinely used within the HCP since 2015, only a handful of studies have focused on its implementation within UK settings from either parent or health professional perspectives [9, 13, 14]. Existing evidence suggested that UK parents found the ASQ-3 acceptable and understandable, however it had the potential to cause anxiety and used Americanised language [7]. Interviews with UK health professionals revealed that they had inconsistent training on the use of the ASQ-3, but that they generally considered it acceptable and understandable, provided some adaptations could be made for an English context [7]. In addition, a report summarising focus groups with parents, health professionals and policymakers at the Department of Health and Social Care identified some aspects of the current tool as needing improvement and questioned its appropriateness for use with children from minoritised ethnic groups and children with disabilities [14]. Survey data from UK health professionals and parents revealed that the purpose of and the responses to the ASQ:SE were not well understood. Health professionals commented on the phrasing of questions and possible responses as being subjective and open to misinterpretation, and parents described certain items as vague or ambiguous [13].

Although the 2-2.5 year visit is mandated, parents uptake of this visit is variable. Analyses of the Community Services Dataset (CSDS), the only national data on health visiting in England, indicated that children living in most deprived areas and looked after children were least likely to have a 2-2.5-year health visitor review recorded, meaning they were also least likely to have a record of the ASQ-3/ASQ:SE [12]. Whilst there were no clear patterns of 2-2.5-year reviews across ethnic groups in the study [12], analyses of the CSDS were limited to local authorities with low levels of missing data, meaning a number of local authorities (such as Bradford, an ethnically diverse and mostly low-income community) were not eligible for inclusion in this study. If ASQ-3 data is missing from more disadvantaged populations within England, this means that the findings from this population level measure may be misleading and may be underestimating the extent of disparities in development by ethnicity and socioeconomic deprivation [15].

Indeed, the qualitative evidence suggests that the ASQ-3 may not be suitable for use with families from some minoritised ethnic groups. Some minoritised ethnic groups may be at heightened risk of poor development due to adverse experiences such as socioeconomic disadvantage, prejudice and racism [16]. Socio-demographic differences could also lower levels of completion in some areas.

Using a subset of the CSDS with 226,505 children at age 2-2.5 years, it was found that whilst 86% of children met expected levels of development overall, inequalities existed with lower rates in areas of high neighbourhood deprivation, some ethnic groups, and in boys [15]. A study completed in ethnically diverse and deprived inner-city areas of Bradford, UK reported lower rates of children reaching expected levels of development, and some indication of variance by socioeconomic characteristics [17].

Since the ASQ-3 is the only routine and nationally recorded measure of child development administered in England before school age, it is crucial to understand if the ASQ-3 is:

a. Acceptable for use with all children and parents - specifically, whether the ASQ is perceived by health professionals as appropriate and satisfactory for its intended purpose of screening and monitoring early child development.
b. Implementable - whether health professionals feel they can successfully deliver the ASQ-3 universally to families as part of their service [18].
c. Equitable - the extent to which the ASQ-3 provides fair and equal opportunities for all children, to access, participate in, and benefit from developmental screening, regardless of ethnicity, socioeconomic status, or other demographic characteristics.

The present study is set within the Born in Bradford’s Better Start (BiBBS) cohort, a sample of 5,700 families living in ethnically diverse and socioeconomically deprived neighbourhoods of Bradford, a city in the North of England [19]. We will address previous evidence gaps by integrating quantitative analysis of the diverse BiBBS cohort data, which includes rich baseline sociodemographic information and linkage to individual-level local health visiting data, with a qualitative exploration of health professionals’ perspectives towards implementing the ASQ-3 and ASQ: SE. We are particularly interested in understanding how these tools perform with families at risk of inequity, including those from minoritised ethnic groups, those who do not speak English or require an interpreter, those living in socioeconomic disadvantage or with a disability (parent or child). The overarching aim is to understand whether the ASQ-3 and ASQ:SE are useful measures of early child development that can be used by health professionals for both clinical practice and research purposes, and if it is suitable to inform policy decision-making.

### Aims and objectives

The overall aim of this mixed-methods study is to investigate if the ASQ-3 and ASQ:SE, delivered via the 0-5 HCP, are acceptable, implementable, and equitable tools for understanding early child development in an ethnically diverse and deprived population.

1. The quantitative component of the study will:

a. Describe how many children receive the mandated 2-2.5-year review from a health professional, and have records for ASQ-3 and ASQ:SE
b. Investigate the sociodemographic factors associated with children receiving a 2-2.5 year review, and having an ASQ-3 and ASQ:SE record
c. Describe differences in the ASQ-3 and ASQ:SE scores by sociodemographic characteristics
2. The qualitative component of the study will explore the perspectives of health professionals on:

a. The implementation of the ASQ-3 and ASQ:SE in practice and whether this differs from relevant guidance
b. The training and support they have received to support the implementation of the ASQ-3 and ASQ:SE in Bradford
c. The barriers and facilitators to implementing the ASQ-3 and ASQ:SE, particularly with families at risk of inequity
d. The purpose, usefulness, and inclusivity of the ASQ-3 and ASQ:SE, particularly for families at risk of inequity
3. The integration component will triangulate: the quantitative findings in objective 1 and the qualitative findings from objective 2, to develop a holistic understanding of how the ASQ-3 and ASQ:SE are being used in practice, and if they are acceptable, implementable, and equitable.

## Materials and Methods

### Study design

This research has a convergent parallel design, where we will use qualitative and quantitative methods independently and then integrate them after analysis through the process of triangulation, see Fig 1 [20]. The benefits of integrating qualitative and quantitative data within a single study are widely recognised. It can enable a deeper and more nuanced understanding of complex phenomena compared with studies using a single method [21, 22]. Within this study context, a mixed-methods approach enables a deeper analysis of the implementation of the ASQs within the Health Visiting Service in Bradford.

**Fig 1.**
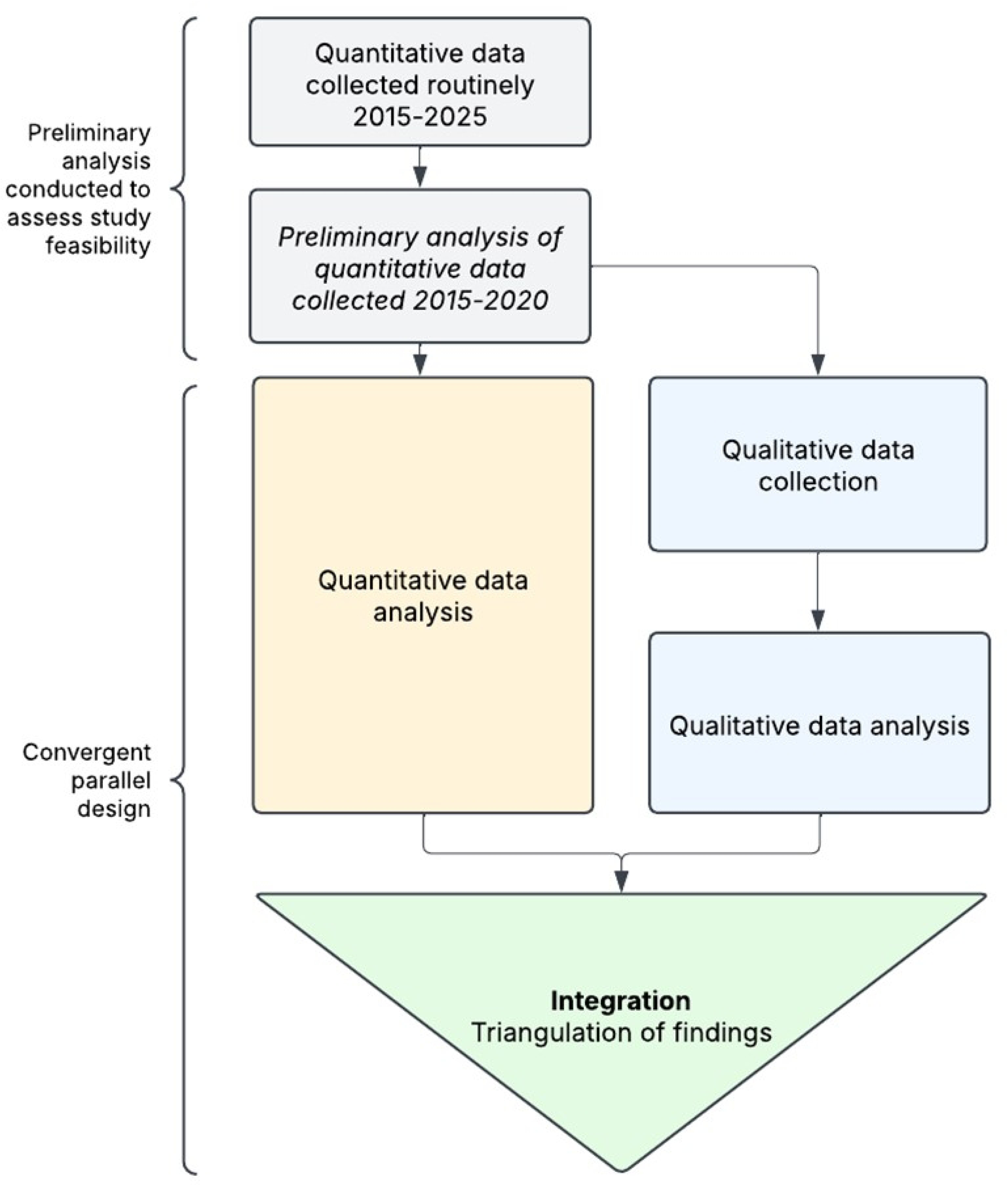
Study overview describing the convergent parallel design.

In this study design, equal weight will be given to both the qualitative and quantitative components. The qualitative and quantitative analyses will be conducted concurrently, and only integrated after the separate analyses are completed.

The quantitative component will use routinely collected health visiting data from a birth cohort study, BiBBS (see setting section below for details). Prior to designing this study, we conducted preliminary quantitative analysis on a subsample of BiBBS participants (those recruited prior to March 2020) to check that an acceptable number of participants’ data could be successfully linked, and ensure the feasibility of the quantitative study for answering our research questions.

### Setting

The setting for this study is Bradford, a city in Northern England with high levels of child poverty, socioeconomic deprivation and ethnic diversity [23].

The quantitative component of this study will be a secondary analysis of data from the BiBBS interventional birth cohort study [19]. BiBBS is a part of the Born in Bradford (BiB) family of studies [24]. BiBBS recruited 5,700 socially disadvantaged families between 2016-2024. All mothers complete an in-depth baseline questionnaire during pregnancy, and consent to routine linkage of their and their child’s routinely collected health and education data. The cohort is representative of the eligible pregnant population, comprising of 88% ethnic minorities, of whom 55% are migrants, 33% have little - no English language and 84% of whom live in the most deprived decile on the Index of Multiple Deprivation [25].

The qualitative component of this study will take place in a single NHS site, Bradford District Care NHS Foundation Trust, and participants will be recruited from the Health Visiting Service. There are four local health visiting teams (East, West, South, and Keighley and Shipley), and each team is made up of a mix of health professionals (e.g., Specialist Community Public Health Nurses (Health Visitors), Staff Nurses, Community Nursery Nurses and Health Visiting Assistants). Health professionals in these teams offer both ASQs (ASQ-3 and ASQ:SE) to all parents in the Bradford District at the 2-2.5 year review.

#### Theoretical Framework

This study will follow a pragmatic approach, drawing on both quantitative and qualitative methods to develop a practical understanding of how the ASQ-3 and ASQ:SE are implemented in Bradford. The Health Equity Implementation Framework (HEIF) will be applied in this study. Implementation frameworks can help understand the determinants that influence the extent to which an intervention is successfully implemented into practice. However traditional implementation frameworks often neglect to consider factors influencing equitable implementation amongst marginalised groups [18]. This is particularly important in this context where we have already identified that inequity exists in administration of the ASQs. HEIF has particular advantages for this study because it:

1. Focuses attention on the clinical encounter and how the interactions during this encounter (in this instance administration of the ASQs) influence implementation. This is important for health disparities as there are unique patient and provider factors that influence healthcare in vulnerable populations.
2. Considers the societal influence on the context, recipient of the ASQ, and the ASQ itself. This includes economies, policies and sociopolitical forces which influence how people live and how healthcare is delivered. In relation to this research, this would include considerations around the setting in which the ASQs are administered, and any wider policies or guidelines influencing its administration.
3. Focuses on health equity determinants related to the recipient of the ASQ, such as lack of trust in the medical system, or low health literacy.

The HEIF will be applied when interpreting the results of both studies to consider how barriers and facilitators to equitable implementation of the ASQ map onto the three healthy equity domains.

### Quantitative component

The quantitative component of this research is an observational secondary analysis of the longitudinal BiBBS cohort. A descriptive epidemiological approach is applied, where we aim to describe the numbers of children that receive the 2-year-visit/ASQs, and the number of children that achieve a Good Level Of Development on these ASQs. We also aim to characterise the distribution of these outcomes by sociodemographic characteristics [26]. We will use the Strengthening the Reporting of Observational Studies in Epidemiology (STROBE) guidelines for reporting our results [27].

#### Eligibility for analyses

Study participants are BiBBS women who:

- Have a completed baseline questionnaire
- Have a child aged at least 3-years-old at the time of analysis (to ensure they would have been eligible for the 2-2.5 year review)
- Have linked BiBBS and health visiting data available at time of analysis.

#### Outcomes

Individual BiBBS pregnancies are linked to routinely collected health visiting activity data within the Bradford District by combining study ID numbers with identifiers such as name, date of birth, address, and/or NHS number. These are shared with the data provider via a secure data transfer method. The data provider then attaches the relevant routine data to the individuals and returns the data via a secure data transfer method. Participant identifiers are removed before the data are returned, leaving only the unique study ID as the link. The linked health visiting dataset includes information on individual health visiting contacts, including whether the appointments were attended, and ASQ records.

From the linked BiBBS health visiting dataset we will derive four binary outcomes: (1) whether parents have a 2-2.5 year visit recorded, (2) whether they have an ASQ-3 and/or ASQ:SE record for their index child at the visit, (3) whether a child met a ‘Good Level of Development’ (GLD) on the ASQ-3 and on the (4) ASQ:SE. These are derived from the following CTV codes: XaQA6 (Health visitor child 24-28 month contact), XacEL-XacEP (ASQ-3 24 month questionnaire scores), XacEQ-XacEU (ASQ-3 27 month questionnaire scores),XacEV-XacEZ (ASQ-3 30 month questionnaire scores), XaaCT, and XafFe (ASQ:SE 24 month and 30 month questionnaire scores).

The ASQs contain three response options to each question (yes/sometimes/not yet, with scores being yes=10, sometimes=5 and not yet=0). Each age version and domain of the ASQ has different cut-offs to categorise children into; (1) ‘below cut-off’, where a child requires further assessment and intervention with a professional, (2) ‘monitor’, where a child’s development should be monitored, and (3) ‘above cut off’, where a child’s development is on schedule. In line with the Public Health England method for calculating whether children have a Good Level of Development (GLD) on an ASQ-3 [28], an overall ASQ-3 and ASQ:SE score will be created by categorising as ‘Not at risk’=above cut off and/or monitor (on all 5 domains for ASQ-3, and overall for ASQ:SE), and ‘At risk’=’below cut off’ (on 1 or more domains for ASQ-3, or overall for ASQ:SE).

#### Predictive measures

To investigate whether the outcomes vary by sociodemographic characteristics, we will use the following measures from the BiBBS baseline questionnaire. We will use two indicators of socioeconomic position. First, maternal education will be categorised into ‘No qualifications’, ‘5 or less GCSEs’, ‘5 or more GCSEs’, ‘A levels’, and ‘Degree’. Second, financial security will be ascertained by mother’s response to “how well are you managing financially?”. Responses will be coded as: ‘Finding it quite/very difficult’, ‘Just about getting by’, ‘Doing alright’, ‘Living comfortably’.

Maternal ethnicity will be grouped into UK Census categories: White British/Irish, British / Asian British Pakistani, White Other (containing Polish, Slovakian, Romanian, Czech, Other White, Gypsy/Roma/Irish Traveller), Other British / Asian British South Asian (containing Indian, Bangladeshi), Black African/Caribbean and Other. The groups may be combined if numbers within ethnic groups are too small and risk reidentification. The other category will not be included in analyses due to its uninterpretable combination of different ethnic groups.

Three further measures will be: maternal country of birth (grouped into Not born in the UK and Born in the UK), maternal first language (grouped into Not English and English), for those whose language is not English, how well they view their English language ability (grouped into not at all/a little/some versus quite/very well), and child sex (grouped into female and male).

#### Analysis

The prevalence of the four outcomes will be described. Binary logistic regression models will be estimated to produce odds ratios and 95% confidence intervals for each outcome. The purpose of the study is to describe differences in the outcomes by sociodemographic characteristics, rather than conduct causal analyses. To do so, we estimate binary logistic regression models by each predictor variable described above, with only analytical adjustment for child age in months at the time of the ASQ-3 as a covariate in the GLD models (as child age may causally influence achievement of a GLD, but cannot be causally related to any of the predictor variables). Adjustment by other variables (e.g. confounders or colliders) could risk misrepresenting the magnitude of the associations by the predictor variables [29, 30]. To compare the magnitude of sociodemographic differences in the outcomes, model-based estimates (ie. adjusted predictions) will be presented.

### Qualitative component

The qualitative component involves semi-structured interviews with health professionals. We have used elements of the COnsolidated criteria for REporting Qualitative research (COREQ) checklist to guide the reporting of the methods in this protocol [31].

#### Sample

A purposive sampling approach will be used to identify and recruit health professionals directly involved in the implementation of ASQs in Bradford and the Better Start Bradford areas. We aim to recruit 20-25 health professionals, with representation from the range of health professionals administering ASQs. Our estimated sample size is informed by considerations of information power, the size of the workforce and capacity to deliver the research. Sampling will be continuously evaluated in weekly research team meetings during the recruitment and data collection period, to help determine whether a sufficient sample size has been achieved to meet the aims and objectives of this research. ‘Information power’ is a concept proposed by Malterud *et al.,* which suggests that the more relevant information the sample holds, the lower the number of participants that are required [32]. It incorporates several elements relevant to determining sufficient information power, including (a) the aim of the study, (b) the sample specificity, (c) the use of established theory, (d) the quality of the discussion, and (e) the analysis strategy. Information power offers a way of ensuring decisions regarding design, sampling, and analysis are robust and defensible.

Inclusion criteria:

a. Health professional (working in an included health visiting team) with direct responsibility for implementing the ASQs as part of the 2.5 year health visiting review or,
b. Team lead or service manager of an included health visiting team

There are no exclusion criteria.

#### Recruitment

The Health Visiting Service will support recruitment by circulating our email invitation to their teams as part of their regular correspondence. The email invitation explains who is eligible to take part in the study and will include a digital copy of the participant information sheet and consent form.

Those interested in taking part will be invited to directly contact the research team using the contact details provided in the participant information sheet and email invitation. A member of the research team will be available to respond to any questions or queries about the research and confirm eligibility. Once eligible participants have agreed to take part in the study, the researcher will arrange a convenient date and time for the interview.

Informed consent will be obtained before proceeding with any interview. Where an interview takes place online or by telephone, informed consent will be confirmed verbally at the beginning of each interview. Verbal consent will be video and/or audio recorded, and the interviewer will also complete and sign a checklist to confirm that this has taken place. Where an interview has taken place face-to-face, informed written consent will be obtained.

#### Data collection

Data collection will be via one-to-one semi-structured interviews with health professionals delivering the ASQ in Bradford, conducted by a member of the research team. All interviewers will be appropriately trained to conduct the interviews and will have supervision and support from senior academics in the research team. To promote accessibility, interviews will take place on either Microsoft Teams, Zoom, by telephone or face-to-face, at the preference of the interviewee.

Each interview is expected to last approximately 60 minutes and will be audio or video recorded. Before the interview begins, participants will be reminded that the interview is being recorded, but that they are able to take part with their camera on or off. Audio or video files will be transcribed using the Microsoft Word transcription function. Transcripts will be anonymised to remove participant-identifiable information, and recordings of the main interview will be deleted once the transcripts have been checked for accuracy.

At the start of the interview, the interviewer will introduce themselves and their links to the project. They will explain the nature and purpose of the interview and how the research outputs will be used and disseminated. Participants will have the opportunity to ask questions throughout.

During the interview, a topic guide will be followed to ensure the key issues are explored with participants (S1 Appendix). The interview topic guide was informed by gaps in existing research, preliminary analysis of BiBBS cohort data and the HEIF. The topic guide will be reviewed after the first few interviews and revised to address any issues of phrasing, although key topics will remain the same. As part of the interview, participants will be shown the ASQ-3 to help them recall specific items and asked for their views on implementing these items in practice. This can add richness to the data by moving from general discussion, into a greater level of specificity [33]. For example, exploring whether there are some questions that do not get answered or are harder to review with parents in certain populations, and if any of the questions or response options are perceived to cause anxiety for parents. We are particularly interested in the application and acceptability of these questions for families at risk of inequity. In efforts to ensure interview timings are accessible to health professionals, the ASQ:SE will not be shown to participants during the interview, however other questions will ask participants to consider both questionnaires. We have decided to discuss ASQ-3 in more detail as it should be used with all children across England, whereas the ASQ:SE is optional in some local areas, however, we will review this after the first few interviews.

At the end of the interview, we will collect information about a participants demographics (e.g., gender, ethnicity, age, first language), as well as information about whether they are a parent, their professional background ( e.g., nursery nurse, school nurse, health visitor), their length of service in total and in Bradford and which area of Bradford they work in.

Researchers conducting qualitative interviews will keep a reflexive journal detailing their overall impression of each interview; the assumptions they made; and their thoughts on how they may have influenced the data collected. These reflexive journal entries will be drawn upon to inform discussions about the analysis.

#### Data analysis

A descriptive thematic analysis will be carried out in six stages, adapted from the approach set out by Braun and Clarke, using NVivo [34, 35]. This approach is well suited to this study as it provides a rigorous but flexible approach to identifying patterns in the data that are relevant to the research objectives. All members of the team will be involved in the analysis.

Researchers who carried out the interviews (LA, HCH) will check the accuracy of the transcripts by reading them whilst listening back to their interview recordings. They will note down their initial thoughts and ideas for codes.

Members of the research team (LA, KM, HCH) will each independently read the same 3 transcripts to familiarise themselves with the data and create a list of initial codes in line with research aims and objectives using open/inductive coding to ensure that unanticipated ideas and findings are captured. A table of codes and illustrative quotes will be shared with the wider research team for discussion, as a coding tree. Transcripts will be coded using the coding tree, with the option to add new codes when needed. Together, the research team will then group codes by meaning and underlying ideas to produce candidate themes and sub-themes around central organising concepts which explain the meaning of the data. These themes will be mapped to the HEIF to explore how the themes link to the different health equity domains.

Themes will be written up and evidenced with a wider set of illustrative quotes and further refined through discussion with the wider research team. The research team will meet regularly during the analysis period to discuss ideas and agree on recurring themes. These themes will also be reviewed with the Head of Community Children’s Services to sense check and consider alternative perspectives on the themes being created.

To report our qualitative findings, we will adhere to the COREQ checklist [31].

### Integration

To integrate the findings, we will adopt the process of triangulation [21]. The findings from each component will be treated with equal importance. The research team will explore where the findings from the three studies converge, offer complementary information, or appear to contradict each other using a ‘convergence coding matrix’ [36]. Researchers will consider whether there is agreement, partial agreement, silence or dissonance between the findings from the two components. This enables researchers to think about the ‘meta themes’ that cut across the findings from the different methods. The HEIF will be used to help interpret the outputs from the integration process.

### Researchers’ positionality

The research team comprises researchers operating in a hospital or university setting and with varying professional experiences in child and early life health. The team members possess expertise in qualitative and quantitative research methods.

The research team is well connected to the Health Visiting Service in Bradford, through setting up this study. Consequently, the research team has some existing understanding about how the Health Visiting Service operates and are aware of some of the current workforce pressures. The research team members conducting both the quantitative and qualitative analyses have knowledge of the preliminary quantitative analyses. We will actively practice reflexivity throughout the research process, meaning we will make a deliberate effort to scrutinize our own beliefs and assumptions and how these have influenced the study design and analysis process. To support this approach, we will challenge each other during our regular study team meetings.

### Data management plan

Bradford Teaching Hospital NHS Foundation Trust (BTHFT) is the data controller for all data arising from the project. The University of York are data processors and will hold and process data for the period of analysis. All data will be stored securely on the University of York Department of Health Sciences secure server and access will be permitted only to authorised members of the research team. No one outside the research team will have access to the data. Once analysis and publications are complete, all data will be transferred for storage and archiving at BTHFT in line with BTHFT information governance policies and existing secure data storage processes. This will allow the de-identified research data to be used for future analyses to advance knowledge in this area. All data and related files will be deleted from the University of York computer systems, and any paper documents either taken to BTHFT or destroyed.

### Patient, Public and Professional Involvement

This research was designed in partnership with the Health Visiting Service in Bradford to ensure the findings are relevant for local practice.

All Better Start Bradford projects and evaluations are supported by a dedicated community research advisory group (CRAG) who meet on a bi-monthly basis. There are nine members of the CRAG, with most being residents in the Better Start Bradford area who were recruited to the group through their involvement in Better Start Bradford projects. The group consists of both mothers and fathers from diverse backgrounds (predominantly South Asian). The CRAG meets to review the acceptability and design of proposed research projects and provide advice on all aspects of the research process. We will work with this group, as well as representatives from the Health Visiting Service, to co-produce the recommendations resulting from our analysis and to plan our dissemination strategy.

### Ethics

The protocol for BiBBS recruitment and collection of routine data was approved by Bradford Leeds NHS Research Ethics Committee (15/YH/0455). Research governance approval was gained from Bradford Teaching Hospitals NHS Foundation Trust. The existing ethics includes approval for observational research using the BiBBS cohort. The current study is thereby covered by the above-mentioned ethical approval. BiBBS takes written informed consent from all participants (ie. parents of children).

The qualitative component of this research was reviewed and approved by the Department of Health Sciences Research Governance Committee at the University of York (*HSRGC/2025/686/A*) and Health Research Authority (HRA) approval was granted (25/HRA/0330).

Any protocol amendments will be authorised by the Chief Investigator and submitted to the NHS sponsor for approval. Standard processes for HRA and research governance committee approvals will be followed.

### Study Timeline

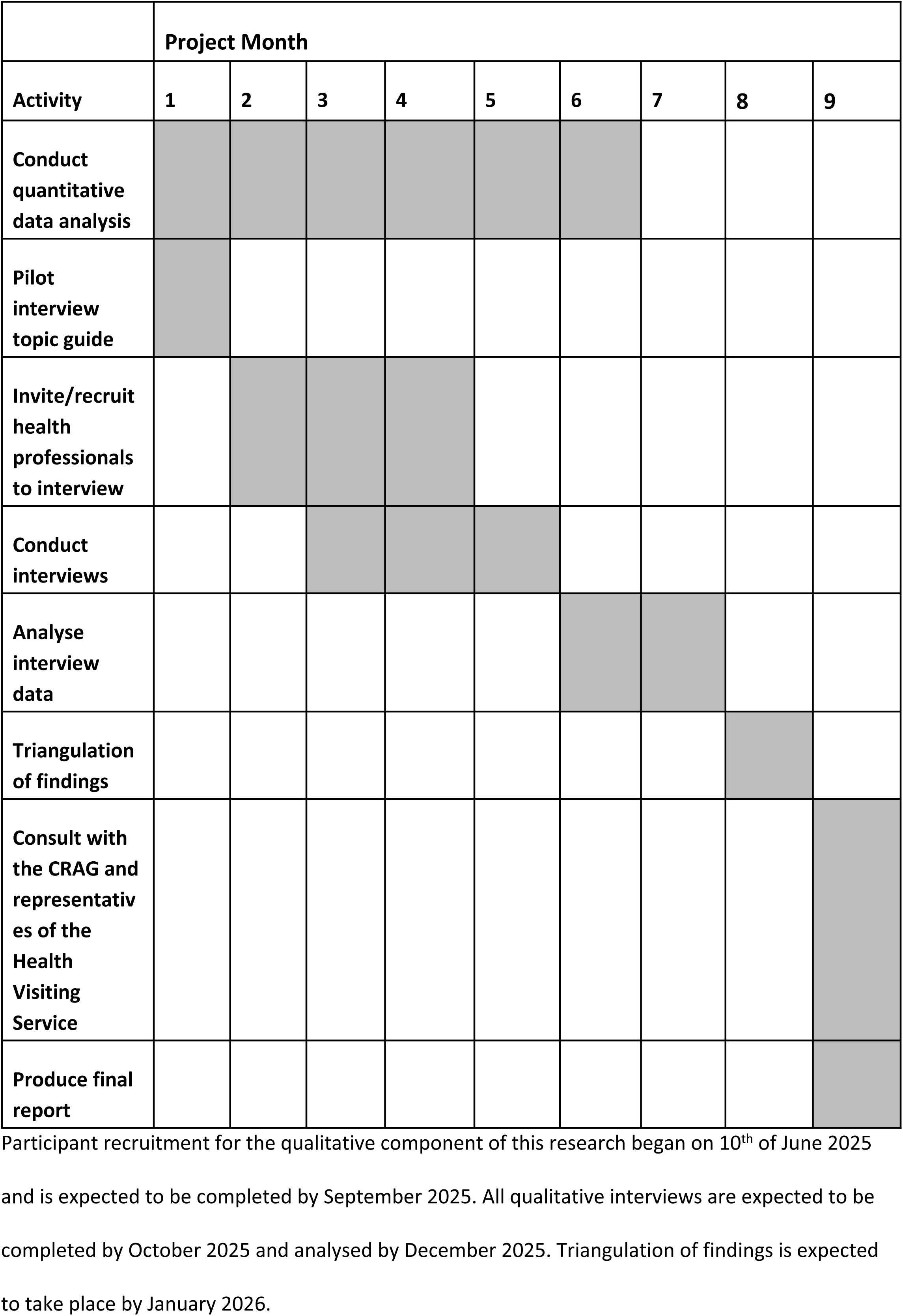

## Discussion

### Contributions

This study aims to understand if the ASQs are acceptable, implementable, and equitable for use by health professionals and families as an overall measure of child development, through a mixed methods study design. The findings from this research will provide crucial insights into the use of the ASQ-3 and ASQ:SE as tools for measuring early child development, particularly in ethnically diverse, urban populations.

If the ASQ-3 and ASQ:SE are found to be acceptable, implementable and equitable, this research will provide confidence in its national use in England. If limitations with using the ASQs are identified, the findings from this research may be used to help improve training for health professionals about implementing the ASQs equitably, overcoming any identified barriers, and how equity in the implementation of ASQs is monitored going forward. For instance, if health professionals report difficulties administering the ASQs in multilingual families and/or minoritised ethnic populations, the study may recommend the development of translated materials and culturally sensitive procedures for collecting ASQ data.

Alternatively, the findings may suggest that an alternative approach to measuring early child development would be beneficial in order to better understand local child developmental needs. For example, a recent rapid literature review identified the Caregiver Reported Early Development Instrument (CREDI) as a potentially feasible alternative to the ASQ-3 [14]. Replacement of the ASQ-3 with an alternative measure of child development would be a significant overhaul to the current system. Therefore, it is important to get clarity on the value of the ASQ-3 before this is considered, particularly given the UK government’s commitment to improving health outcomes for all babies [37], which underlines the importance of acceptable and equitable screening in early childhood.

This research can also inform whether the ASQ-3 and ASQ:SE can be confidently utilised as a research tool to investigate child development in cohort studies that link routinely collected data (e.g. BiBBS, ECHILD, BiB4All) [19, 38, 39]. Members of our team plan to use this data to investigate inequalities in children’s outcomes and to evaluate early years interventions. This is important, given the impact of early development delays on outcomes later in life if not detected and addressed, and the mandated use of the ASQ-3 at the 2-2.5 year health visitor review in England.

In conclusion, this research will provide valuable insights into the acceptability and implementation of the ASQs in an urban, ethnically diverse, socioeconomically disadvantaged setting. By combining quantitative data regarding the disparities in receiving a 2-2.5-year review and an ASQ, with in-depth interviews with health professionals using the ASQs, this study will contribute to the delivery of more equitable early child development services in Bradford and beyond.

### Strengths and limitations of the study

This research will address key evidence gaps regarding the acceptability of implementing the ASQ-3/ASQ:SE in a UK setting, with a particular focus on diverse populations. This study is set in Bradford and has been designed with support from the local Health Visiting Service in Bradford. By providing evidence-based recommendations for improving the implementation of the ASQs, this research will contribute to the delivery of more effective and equitable services in the local area. Moreover, the study will generate data that can be used to support applications for further funding, enabling the service to expand its capacity and reach.

Previous research into child development in England has used ASQ-3 data captured in CSDS [15]. ASQ-3 data are entered into each local data system by providers of health visiting and then uploaded monthly to the CSDS, where it is collated at a national level. In this study, we used rich baseline questionnaire data collected for BiBBS participants and linked directly to the individual-level ASQ-3 and ASQ:SE data captured in the local data system used by Health Visitors. This offered the opportunity to use the most up to date information about families in Bradford, to understand how this is captured at a local level.

The BiBBS cohort recruited some of the most socially disadvantaged families in England, comprising 88% ethnic minorities, and 55% migrants, with 56% with maternal obesity, 46% experiencing poor maternal mental health, and 23% reporting financial insecurity [25]. This study will therefore provide evidence that can be generalised to similarly populated urban inner-city areas, including families who may be in most need of support.

A limitation of the quantitative work is that it will not be possible to disentangle the difference between truly missing data and data that has not been input into the health visiting records, hence we may overestimate the amount of missing ASQs in our study. However, the study will still provide insights into which children are most or least likely to have ASQ records.

Currently, there are a number of national challenges facing the Health Visiting Service, including funding cuts and workforce shortages. In a recent survey by the Institute of Health Visiting, a large percentage of health visitors (74%) said that workforce shortages impacted on the delivery of universal health visiting contacts and 73% said this impacted on their ability to support families in need [40]. Since 2015, England has lost more than 40% of its health visitors meaning that many families are missing out on vital support. Given these workforce pressures, this could make it challenging to implement any changes identified in this study. The Health Visiting Service in Bradford have been part of the development of this protocol and are supportive of this work, which we hope will facilitate the implementation of the recommendations following this research.

### Dissemination plan

Findings from the study will be written up for publication in peer-reviewed scientific journals and as conference abstracts, and shared amongst relevant partners, including local and national stakeholders and commissioners. A summary of the findings will also be disseminated to the community via the CRAG and Better Start Bradford Innovation Hub website (https://borninbradford.nhs.uk/what-we-do/improving-health/bsb-innovation-hub/). A copy of the summary and the final report can be made available to the qualitative research participants.

Participants can confirm if they would like to receive the summary report when they consent to take part in the research. No participants will be identifiable in any reports or publications produced from this work. Participants can also request a copy of any publications by contacting the research team using the contact details on the participant information sheet.

## Data Availability

No datasets were generated or analysed during the current study. All relevant data from this study will be made available upon study completion.

## Acknowledgements

The integration of research and practice in Bradford has only been possible because of the enthusiasm and commitment of staff and volunteers across children’s services in Bradford. We are grateful to all Born in Bradford staff, the Better Start Bradford partnership and staff, all Better Start Bradford project teams, health professionals, local authority and voluntary and community sector organisations who have supported the integration of research into practice. We are grateful to all the families taking part in BiBBS and all members of the Community Research Advisory Group.

## Supporting Information

S1 Appendix: Interview Topic Guide

## Notes

### Competing Interest Statement

The authors have declared no competing interest.

### Author Declarations

The protocol for BiBBS recruitment and collection of routine data was approved by Bradford Leeds NHS Research Ethics Committee (15/YH/0455). Research governance approval was gained from Bradford Teaching Hospitals NHS Foundation Trust. The existing ethics includes approval for observational research using the BiBBS cohort. The current study is thereby covered by the above-mentioned ethical approval. BiBBS takes written informed consent from all participants (ie. parents of children). The qualitative component of this research was reviewed and approved by the Department of Health Sciences Research Governance Committee at the University of York (HSRGC/2025/686/A) and Health Research Authority (HRA) approval was granted (25/HRA/0330). Informed written or verbal consent will be obtained for participants of the qualitative component of this research.

